# Pre-lung transplant monocyte counts predict post-lung transplant survival and adverse outcomes in IPF

**DOI:** 10.1101/2025.05.26.25328338

**Authors:** Theodoros Karampitsakos, Muhammad Raheel Qureshi, Jalen Hammonds, Christian Arce Guzman, Rebecca Albuquerque, Bochra Tourki, Zainab Fatima, Nicole Henriquez, Valeria Calderon, Tamer Fadli, Amanda McNamara, Ishna Poojary-Hohman, Brenda M Juan-Guardela, Debabrata Bandyopadhyay, Kapil Patel, Jose D. Herazo-Maya

## Abstract

**Introduction:** Accurate pre-lung transplant biomarkers of post-lung transplant survival are lacking in Idiopathic Pulmonary Fibrosis (IPF).

**Methods:** This was a retrospective, observational study including consecutive patients diagnosed with IPF at the University of South Florida/ Tampa General Hospital. First, we compared survival differences in patients with IPF that received lung transplant versus non-recipients, then we investigated whether pre-transplant monocyte counts could predict post-lung transplant survival, Primary Graft Dysfunction (PGD), Acute Cellular Rejection (ACR), Antibody-Mediated Rejection (AMR) and Chronic Lung Allograft Dysfunction (CLAD) using Cox Proportional Hazards (CoxPH) models adjusted to Gender, Age and Physiology index (GAP).

**Results:** A total of 201 patients with IPF were included in the analysis [lung transplant recipients: n=103, non-recipients of lung transplant: n=98]. Patients with IPF that did not undergo lung transplantation had significantly worse survival compared to patients with IPF that underwent lung transplantation [3.13 years (95% CI: 2.30 to 3.72) vs 7.05 years (95% CI: 5.41 to 8.48), HR: 2.95 (95% CI: 2.18 to 4.00), p<0.0001]. Patients with IPF and pre-lung transplant monocyte counts>700 K/μL had increased risk of post-lung transplant mortality [HR: 1.71 (95%CI: 1.10 to 2.65), p=0.016] or adverse outcomes defined as either PGD, ACR, AMR or CLAD, [HR: 2.05 (95% CI: 1.11 to 3.78), p=0.02] compared to patients with monocyte counts≤700 K/μL.

**Conclusion:** Lung transplantation substantially prolongs survival of patients with IPF. Incorporation of pre-lung transplant monocyte counts in the pre-transplant evaluation of patients with IPF could optimize the selection of ideal lung transplant candidates with increased probability of survival.

## Introduction

Idiopathic Pulmonary Fibrosis (IPF) remains a chronic lung disease with dismal prognosis despite the advent of antifibrotic compounds(1). The dismal prognosis of IPF and the concomitant advent of life-extending options in other chronic lung diseases, such as cystic fibrosis(2), is leading to a constantly increasing proportion of adult lung transplants performed for IPF(3–5). In fact, lung transplantation for various end-stage forms of fibrotic interstitial lung diseases has emerged as the most common indication for lung transplantation(3, 4, 6). In the context of IPF, some reports with moderate sample size, showed that lung transplantation can improve survival, mainly through comparison of transplanted patients versus those on waiting list(7–9). However, estimation of survival benefit in larger IPF cohorts and identification of accurate pre-transplant biomarkers of post-transplant survival in IPF represent unmet needs. With donor pool being too small to meet demand and with mortality on transplant waiting lists remaining high, accurate pre-transplant biomarkers might revolutionize the management of such patients and reduce disparities(5, 10).

IPF is thought to be a lung limited disease that does not recur after lung transplantation, however, the fact that circulating, myeloid-derived monocytes and monocyte-expressed genes predict IPF mortality and disease progression, suggests that a systemic component cannot be entirely excluded in IPF(11–15). Since the myeloid compartment remains unchanged after lung transplantation in IPF and since most immunosuppressive agents used after lung transplant do not target monocytes directly, we hypothesized that increased monocyte counts pre-lung transplant in IPF could be associated with increased mortality risk and other adverse outcomes after lung transplant and serve as a potential biomarker to identify optimal candidates for lung transplantation.

Our single-center study confirmed that lung transplantation substantially prolongs survival of patients with IPF and identified a pre-lung transplant absolute monocyte count of 700 K/μL as the ideal cutoff to stratify patients with increased risk of mortality and adverse outcomes post-lung transplantation in IPF.

## Methods

### Study design and participants

This was a retrospective, observational study including consecutive, patients with IPF. Patients with IPF referred to Tampa General Hospital/ University of South Florida between 8/2/2011 and 10/22/2020 were included in the analysis. Diagnosis of IPF was based on American Thoracic Society/ European Respiratory Society/ Japanese Respiratory Society/ Latin American Thoracic Association guidelines (16, 17).

### Pre-lung transplant and post-lung transplant assessment

Following pharmacological and non-pharmacological treatment for IPF, patients were referred for pre-transplant evaluation, typically at the time point that they were in need of 2 liters per minute supplemental oxygen at rest or if Modified Medical Research Council Dyspnea Scale was 3 or 4. Patients underwent pre-transplant assessment at Tampa General Hospital and in case of no absolute contraindication (and preferably if Body-Mass Index<32 kg/m^2^ ) received single lung or bilateral lung transplantation. When the organs were available, organ allocation was decided by a senior member of the transplant team. Routine medical management was offered to all patients during the waiting period under the supervision of a senior transplant physician. Following lung transplantation, patients were examined at regular intervals at the outpatient clinic of Tampa General Hospital and were closely monitored for unusual symptoms or functional impairment.

### Data collection

Data for patients including time of IPF diagnosis, age, gender, forced vital capacity (FVC) %predicted, diffusing capacity of the lung for carbon monoxide (DLCO) %predicted, monocyte counts within 2 months pre-lung transplant, comorbidities, medications prescribed, post-lung transplant treatment-related adverse events, Primary Graft Dysfunction (PGD), Antibody-Mediated Rejection (AMR), Acute Cellular Rejection (ACR) or Chronic Lung Allograft Dysfunction (CLAD), time and type of lung transplantation, time of death were extracted from EPIC. Monocyte counts selected for analysis were the closest to lung transplant.

### Objectives

The objectives of the study were:

1. To investigate survival differences between patients with IPF that were recipients and non-recipients of lung transplant.
2. To investigate if pre-lung transplant monocyte counts can predict post-lung transplant survival.
3. To investigate if pre-lung transplant monocyte counts can predict post-lung transplant adverse outcomes including PGD, AMR, ACR and CLAD.

### Statistical analysis

Summary descriptive statistics were generated with categorical data displayed as absolute numbers and relative frequencies. Continuous data were denoted as mean ± standard deviation (SD) or medians with 95% confidence interval (95% CI) based on presence or absence of normality following the Kolmogorov-Smirnov test. We investigated survival differences from the time point of IPF diagnosis between recipients and non-recipients of lung transplant. We performed both univariate analysis and multivariate Cox-regression models adjusted to IPF-Gender, Age, Physiology (GAP) score at diagnosis (18).

We also performed a subgroup analysis in the recipients of lung-transplant and investigated differences in post-lung transplant survival based on the pre-lung transplant monocyte counts. In particular, we extracted monocyte counts from the complete blood count obtained within the last 2 months before lung transplantation. Receiver operating characteristic (ROC) curve was used to identify the optimal threshold of the pre-transplant monocyte counts for 5-year post-lung transplant mortality prediction. Patients were split into two groups based on that threshold. Except overall survival, we studied 5-year survival because it’s a widely used metric for the evaluation of post-lung transplant outcomes(19–21). Survival differences were presented based on the Kaplan-Meier method. We performed both univariate analysis and multivariate Cox-regression models adjusted to pre-lung transplant GAP score(18). Similarly, we investigated if patients with pre-transplant monocyte counts above this threshold had increased risk for the composite endpoint of PGD, AMR, ACR or CLAD.

### Ethics approval

Ethical approval for this study was given by the Institutional Review Boards / Research Integrity & Compliance of University of South Florida (STUDY: 008038).

## Results

### Baseline characteristics

A total of 201 patients with IPF were included in the analysis [lung transplant recipients: n=103, non-recipients of lung transplantation: n=98]. Overall, most patients were males (n=141, 70.1%) and median age at the time point of referral was 64.51 years (95% CI: 63.57 to 65.70). Baseline characteristics for recipients and non-recipients of lung transplant are presented in **Table 1** . Functional indices at referral were comparable between groups. Median FVC% predicted was 61.0 (53.90 to 73.0) and 64.0 (54.0 to 69.24) in recipients and non-recipients of lung transplant, respectively (p=0.37). Median DLCO% predicted was 41.0 (34.0 to 48.0) and 38.0 (36.0 to 41.62) in patients that underwent and did not undergo lung transplantation, respectively (p=0.59). Mean GAP at the time point of diagnosis was 4.3±1.4 and 4.5±1.3 in recipients and non-recipients of lung transplant, respectively (p=0.21).

**Table 1.** Demographics and characteristics of patients.

| Characteristics | Lung Transplant<br>Recipients<br>(N, %) | No Lung Transplant<br>Recipients<br>(N, %) |
| --- | --- | --- |
| Number of patients | 103 | 98 |
| Male gender/Female gender | 78 (75.7%)/25(24.3%) | 63(64.3%)/ 35(35.7%) |
| Age at referral (median, 95% CI) | 63.44 (62.40 to 64.50) | 66.40 (64.50 to 68.64) |
| Current smokers | 0 (0%) | 6 (6.12%) |
| Ex smokers | 72 (69.90%) | 68 (69.39%) |
| Never smokers | 31 (30.10%) | 24 (24.49%) |
| FVC% pred (median, 95% CI) | 61.0 (53.90 to 73.0) | 64.0 (54.0 to 69.24) |
| DLCO% pred (median, 95% CI) | 41.0 (34.0 to 48.0) | 38.0 (36.0 to 41.62) |
| Antifibrotic treatment | 48 (46.66%) | 60 (61.22%) |
| GERD | 95 (92.23%) | 62 (63.27%) |
| Arterial hypertension | 84 (81.55%) | 67 (68.37%) |
| Dyslipidemia | 83 (80.58%) | 72 (73.47%) |
| Chronic heart disease | 73 (70.87%) | 53 (54.08%) |
| Pulmonary hypertension | 67 (65.05%) | 60 (61.22%) |
| Diabetes mellitus | 42 (40.78%) | 39 (42.86%) |
| Hypothyroidism | 25 (24.27%) | 18 (18.37%) |

**Abbreviations:** CI: Confidence Interval, FVC: Forced Vital Capacity, DLCO: Diffusing capacity of the lung for carbon monoxide, GERD: Gastroesophageal Reflux Disease

### Lung transplantation improves survival in patients with IPF

A total of 103 patients with IPF underwent lung transplantation [bilateral lung transplantation: 65/103 (63.1%), single: 38 (36.9%)]. Patients with IPF that did not undergo lung transplantation had significantly worse overall survival compared to patients with IPF that underwent lung transplantation [median survival from diagnosis: 3.13 years (95% CI: 2.30 to 3.72) vs 7.05 years (95% CI: 5.41 to 8.48), HR: 3.26 (95% CI: 2.36 to 4.50), p<0.0001], **(Figure 1A)**. Increased mortality risk for non-recipients of lung transplant was also observed following Cox regression adjusted to GAP [HR: 2.95 (95% CI: 2.18 to 4.00), p<0.0001], **(Figure 1B)**.

**Figure 1.**
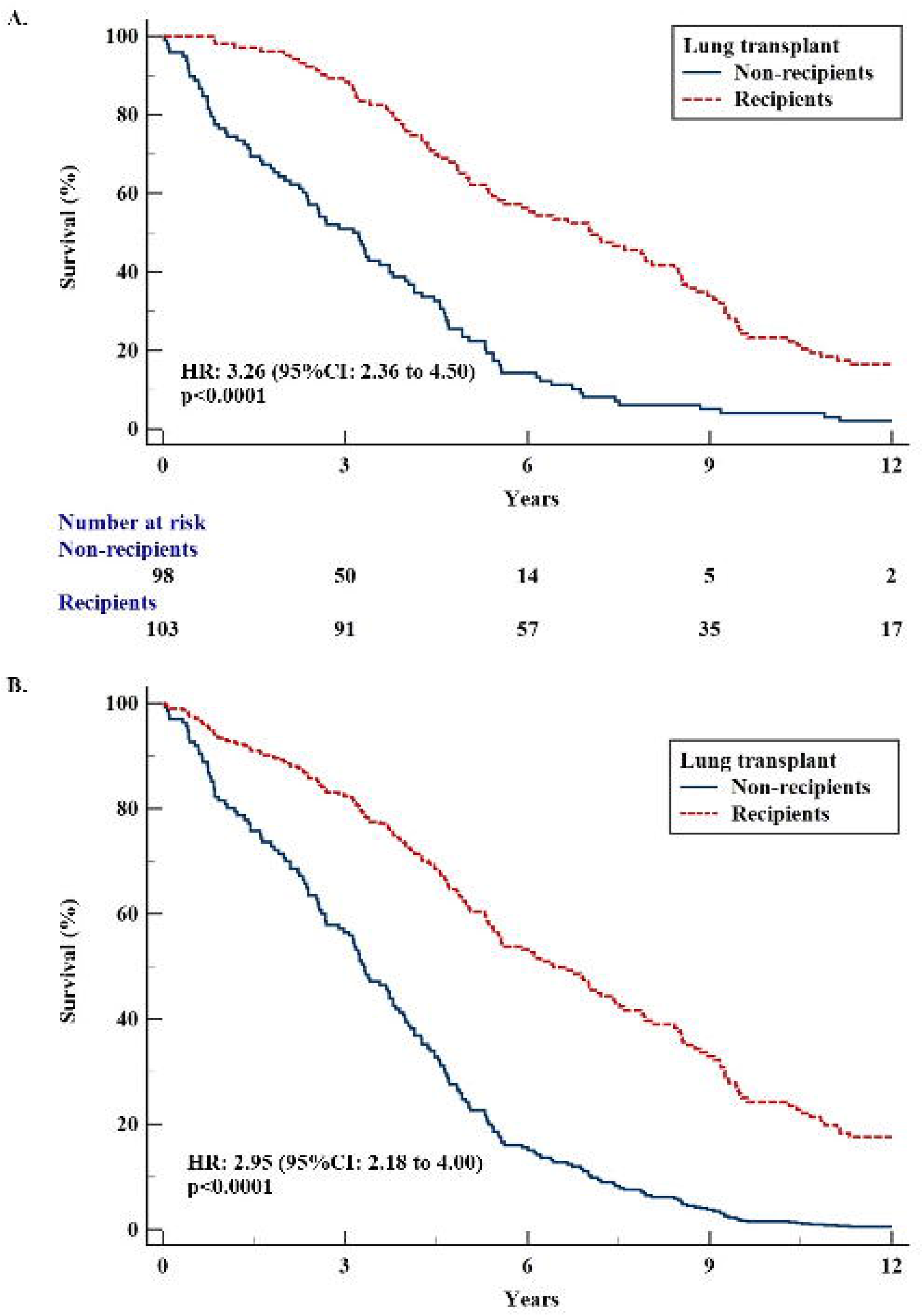
Survival is significantly different in patients with IPF that underwent lung transplantation compared to those that did not both in the univariate analysis (Panel A) and in the Cox-regression adjusted to GAP (Panel B).

### Pre-lung transplant monocyte counts predict post-lung transplant survival

Pre-lung transplant monocyte counts were available in EPIC for 85 patients with IPF that underwent lung transplantation (85/103, 82.5%). Mean value of pre-lung transplant monocyte counts was 776 K/μL ± 270. We performed ROC for the 5-year post-lung transplant mortality prediction based on existing literature using this metric(19–21) and identified the value>700 K/μL as the optimal threshold of pre-lung transplant monocyte count for this endpoint. Patients with IPF and pre-lung transplant monocyte counts>700 K/μL had increased risk for 5-year post-lung transplant mortality compared to patients with monocyte counts≤700 K/μL [univariate analysis – HR: 1.91 (95%CI: 1.16 to 3.14), p=0.011, Cox regression adjusted to GAP - HR: 1.94 (95%CI: 1.15 to 3.25), p=0.012], **(Figure 2A, B)** . The prognostic accuracy was reproducible in overall mortality. Patients with IPF and pre-lung transplant monocyte counts>700 K/μL had increased risk for post-lung transplant all-cause mortality [univariate analysis – HR: 1.72 (95%CI: 1.11 to 2.66), p=0.015, Cox regression adjusted to GAP - HR: 1.71 (95%CI: 1.10 to 2.65), p=0.016], **(Figure 2C, D)**.

**Figure 2.**
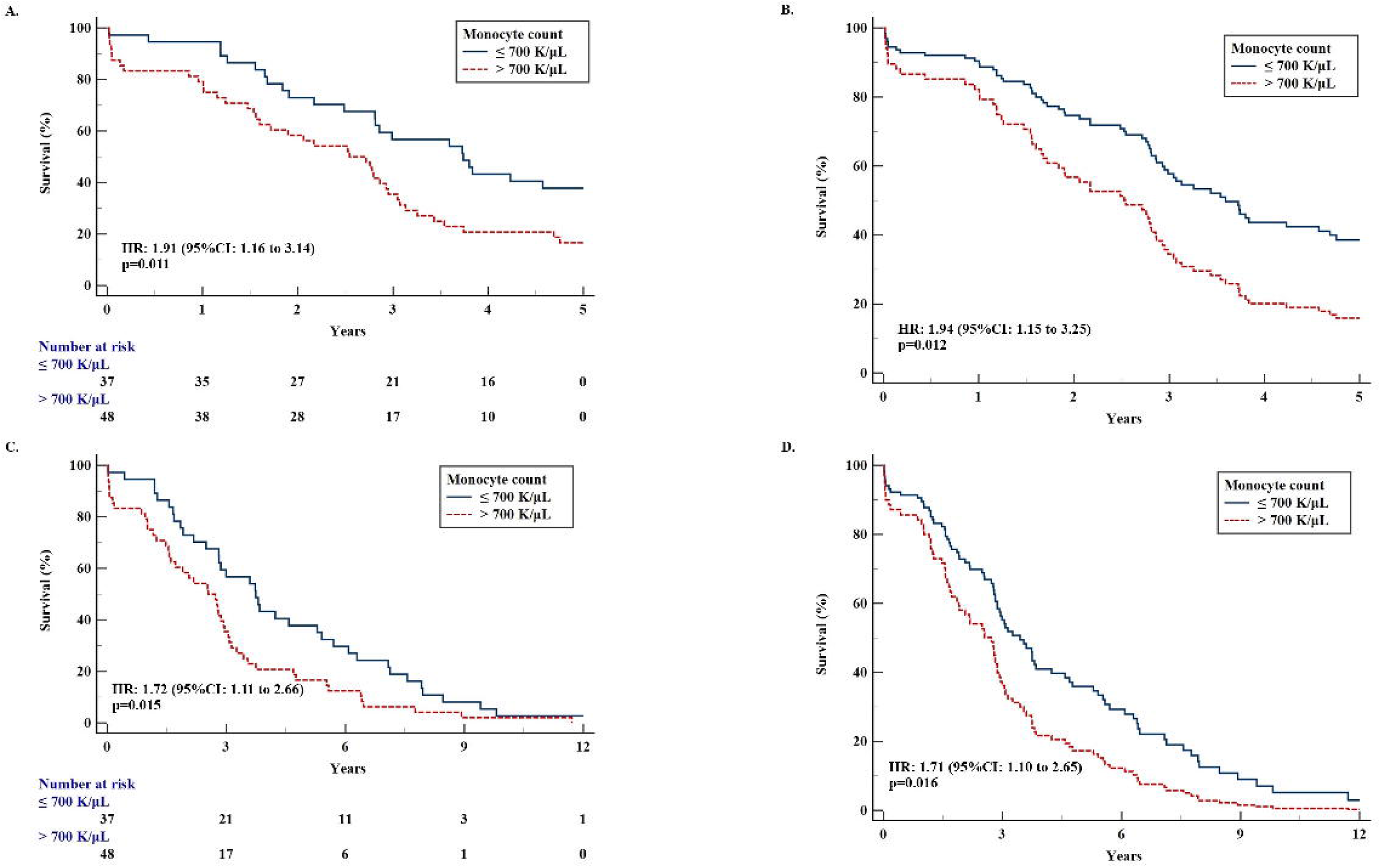
Post-lung transplant 5-year survival is significantly different in patients with IPF and pre-lung transplant monocyte counts>700 K/μL compared to those with ≤ 700 K/μL both in the univariate analysis (Panel A) and in the Cox-regression adjusted to GAP (Panel B). Similarly, post-lung transplant overall survival is significantly different in patients with IPF and pre-lung transplant monocyte counts>700 K/μL compared to those with ≤ 700 K/μL both in the univariate analysis (Panel C) and in the Cox-regression adjusted to GAP (Panel D).

### Pre-lung transplant monocyte counts predict post-lung transplant adverse outcomes

Treatment following lung transplantation is presented in **Table2** . Proportions represent the percentage of patients that received this compound at any time point after lung transplantation. Most patients received corticosteroids (103/103, 100%), tacrolimus (99/103, 96.1%) and mycophenolate mofetil (95/103, 92.2%). Adverse events are presented in **Table 3**, with most common being hyperglycemia (90/103, 87.4%). Data for lung transplant rejection are presented in **Figure 3A**. PGD occurred in 17.5% of the cohort (18/103), AMR in 10.7 % of patients (11/103), ACR in 33.0% of patients (34/103), CLAD in 35.0% of patients (36/103), while any of the aforementioned occurred in 62.1% of the patients (64/103). Importantly, the same threshold of pre-lung transplant monocyte counts that split patients into two groups with significant differences in post-lung transplant survival, split also patients into two groups with significant differences in the risk for PGD, AMR, ACR or CLAD development [Cox regression adjusted to GAP - HR: 2.05 (95% CI: 1.11 to 3.78), p=0.02, **Figure 3B**].

**Figure 3.**
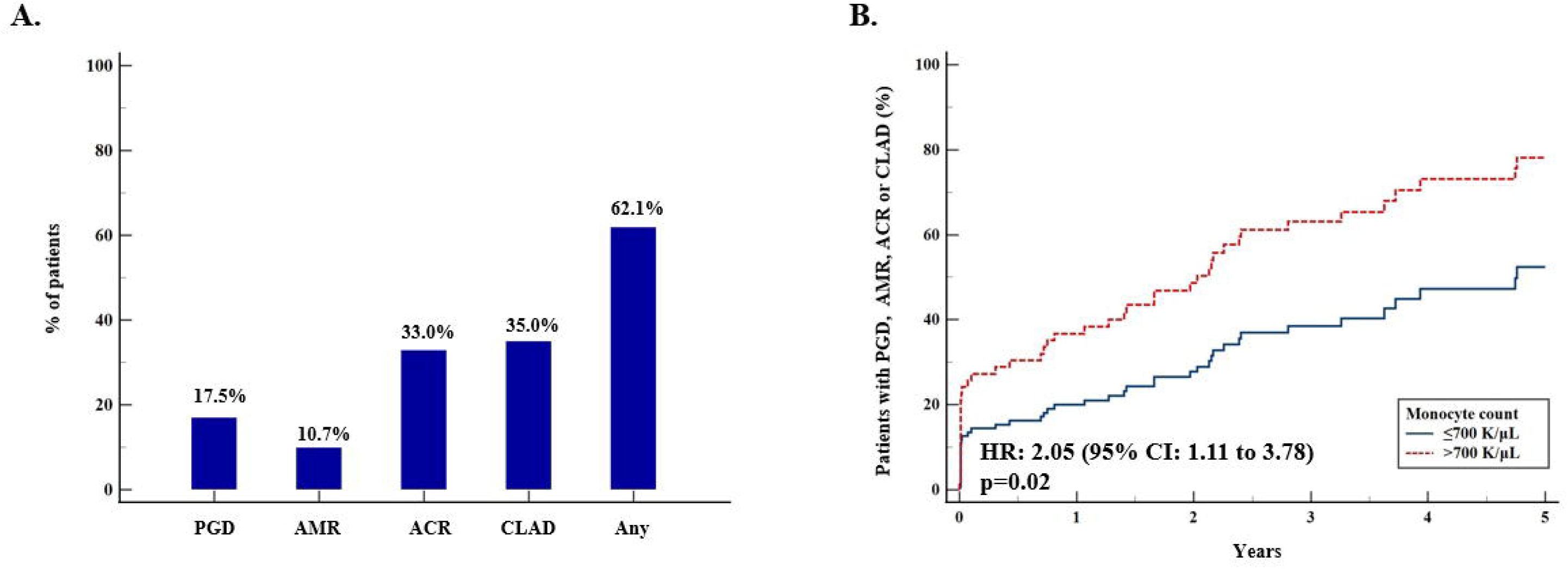
Proportion of patients that experience PGD, AMR, ACR or CLAD (Panel A). Of note, the same threshold of pre-lung transplant monocyte counts that split patients into two groups with significant differences in post-lung transplant survival, split also patients into two groups with significant differences in the risk for PGD, AMR, ACR or CLAD development [Cox regression adjusted to GAP - HR: 2.05 (95% CI: 1.11 to 3.78), p=0.02, (Panel B)].

**Table 2.** Post-lung transplant treatment.

| Compound | (N, %) |
| --- | --- |
| Corticosteroids | 103 (100%) |
| Tacrolimus | 99 (96.11%) |
| Mycophenolate mofetil | 95 (92.23%) |
| Basiliximab | 64 (62.13%) |
| Azathioprine | 41 (39.80%) |
| Sirolimus | 23 (22.33%) |
| Cyclosporine | 20 (19.41%) |
| Rituximab | 4 (3.88%) |

**Table 3.**
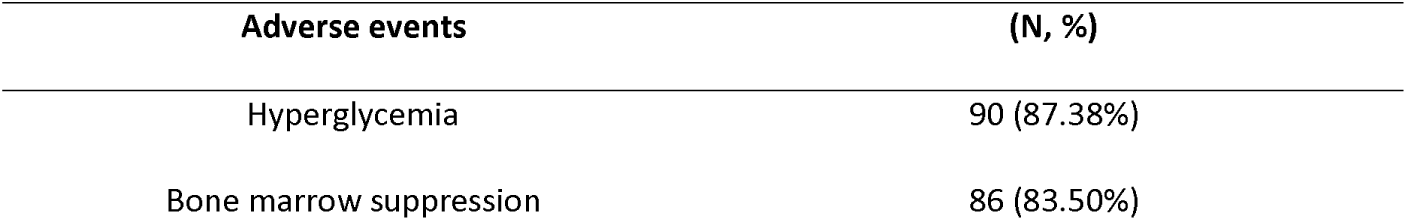
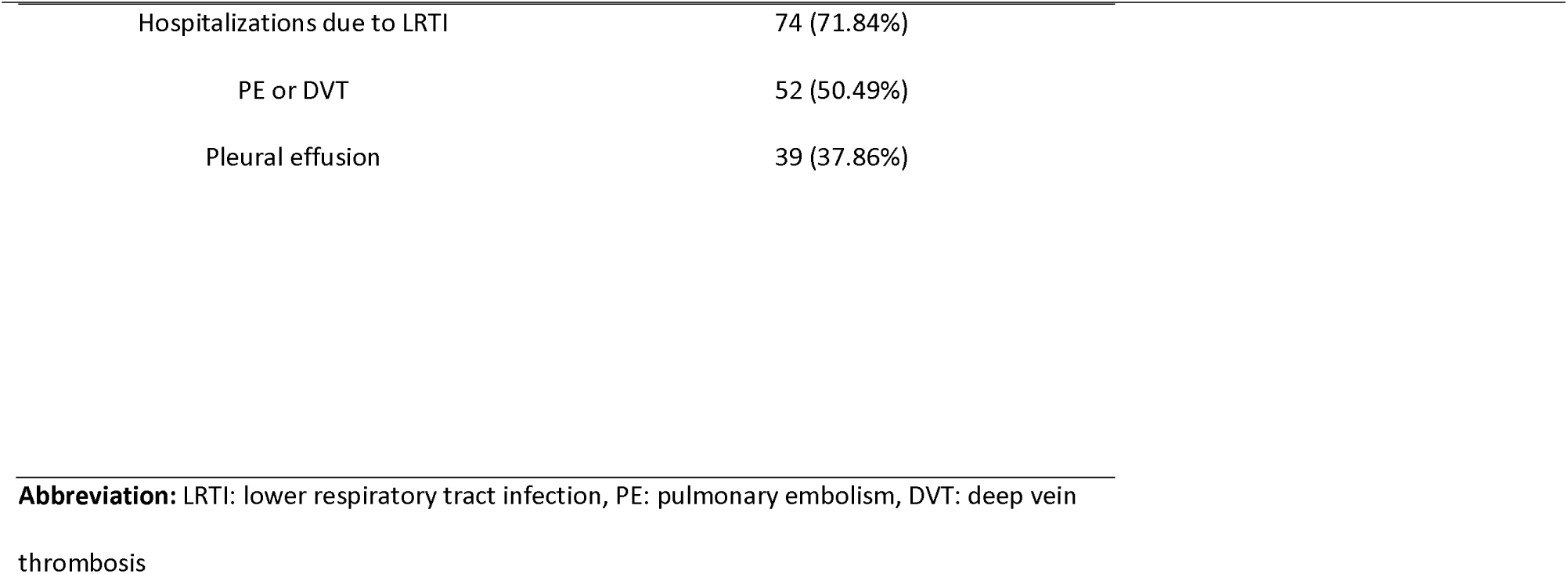
Post-lung transplant treatment related adverse events.

| Adverse events | (N, %) |
| --- | --- |
| Hyperglycemia | 90 (87.38%) |
| Bone marrow suppression | 86 (83.50%) |
| Hospitalizations due to LRTI | 74 (71.84%) |
| PE or DVT | 52 (50.49%) |
| Pleural effusion | 39 (37.86%) |

**Abbreviation:** LRTI: lower respiratory tract infection, PE: pulmonary embolism, DVT: deep vein
thrombosis

## Discussion

This study highlights the survival benefit that lung transplantation confers to patients with IPF and provides a potential novel biomarker for the pre-lung transplant evaluation of patients with IPF. We analyzed survival data of patients with IPF and demonstrated that recipients of lung transplant exhibited a significant survival benefit compared to non-recipients. Importantly, we identified pre-lung transplant monocyte counts as a biomarker able to predict post-lung transplant survival. Pre-lung transplant monocyte counts were also able to predict the risk for the adverse outcome composite endpoint of PGD, AMR, ACR or CLAD, implying that monocytes might have a key role in either acute or chronic transplant rejection. These findings could optimize the selection of ideal lung transplant candidates with increased probability of survival. These data also might imply that IPF is not a ‘’lung- limited’’ disease given that even after lung transplantation patients with higher monocyte counts have worse prognosis. The main attributes of this work are discussed in detail below.

First, a main attribute of this work is that we present survival differences between recipients and non-recipients of lung transplant from the time point of IPF diagnosis in a relatively large cohort compared to existing literature. That was an unmet need and might unveil the ‘’true’’ benefit from lung transplantation in IPF. In particular, a previous study included 46 patients with IPF that were accepted for lung transplantation and used Cox proportional-hazards models to compare those that underwent lung transplantation with those on a waiting list (7). This analysis showed a survival benefit for the patients that underwent lung transplantation(7). A more recent study with a similar approach also demonstrated survival benefit following lung transplantation and suggested that the benefit is sustained for patients with more than 65 years of age(9). Despite that these data were promising, comparison of patients that underwent lung transplantation versus those on the waiting list might not be totally representative, because these groups of patients might not be really comparable due to differences in factors such as frailty and life expectancy. Similarly, comparison of survival between patients with IPF that underwent lung transplantation and a historical cohort has limitations(22). Other multivariate models using lung transplantation as a time-dependent covariate, multiphase hazard models or studies analyzing various interstitial lung diseases (ILDs) have also been reported(6, 8, 23–26). However, given that a randomized controlled trial between recipients and non-recipients does not seem ethical, our approach seems to be an important addition to the literature. Patients of this cohort were followed up from the time point of diagnosis, were treated using a homogeneous algorithm after the referral to our Hospital and our analysis suggested that patients with IPF that undergo lung transplantation have an overall median survival of over 7 years.

Second, another very important attribute of this work is the identification of a biomarker that is tied to the pathogenesis of IPF, as a biomarker able to contribute in the pre-lung transplant evaluation of patients with IPF. Pre-lung transplant monocyte counts predicted post-lung transplant survival even after adjustment to GAP score. We tested both 5-year survival and overall survival post-lung transplant, given that 5-year post-transplant mortality is a widely used metric in post-lung transplant follow-up(19–21). Our findings couple with previous findings for monocyte-specific genes and monocytes in stable IPF(27, 28). In particular, a 52-gene signature in peripheral blood was able to predict mortality in IPF in six independent cohorts (11, 12). Cellular deconvolution of gene expression data showed that monocytes were the cellular source of the upregulated genes(13, 29, 30). This fueled large-scale studies which demonstrated that increased monocyte counts were associated with increased risk of disease progression, hospitalization and mortality in IPF(13–15). Extensive research effort suggested that monocytes do not serve only as biomarkers, but also participate in the pathogenesis of IPF(31–33). The finding that pre-lung transplant monocyte counts predict outcomes even after lung transplant leads to an open question for a potential systemic component as a driver of lung fibrosis progression. Moreover, the findings of the current work extend the prognostic role of monocytes in the pre-lung transplant evaluation and could fuel mechanistic studies aiming to address whether monocytes have a role in lung transplant rejection. Besides, the fact that pre-lung transplant monocyte counts predicted the risk for PGD, AMR, ACR or CLAD suggests that monocytes might have a key role in transplant rejection.

Currently, prognostication of post-lung transplant IPF survival is largely based on non-disease specific factors including Lung Allocation Score, comorbidities such as pulmonary hypertension, esophageal dysmotility and Body Mass Index(6, 25, 34, 35). While the aforementioned have a cardinal role in the pre-transplant evaluation of patients with IPF, coupling them with disease-specific biomarkers such as pre-transplant monocyte counts might revolutionize pre-transplant evaluation by optimizing the selection of ideal lung transplant candidates or by highlighting the need of meticulous evaluation is some recipients of lung transplant. With an increasing proportion of transplants being used for patients ILD, it is important to increase the awareness for the complexities related particularly to this population and shift towards a personalized management approach(6, 27, 28). Towards this direction, other biomarkers, tied to the pathogenesis of pulmonary fibrosis, such as telomere length have been investigated as potential prognosticators of post-lung-transplant outcomes in IPF(8, 36, 37).

Despite the importance of our findings, we need to recognize some of the limitations of our study. First, this was a single-center study. While validation of the absolute monocyte count cut-off identified by us in an additional cohort would be required before using monocyte counts in the pre-transplant evaluation of IPF patients, we strongly believe that a cohort of more than 100 transplanted patients with IPF such as ours, is an adequate sample size given the existing literature for lung transplantation in IPF. Second, our study has the inherent weakness of a retrospective study.

In conclusion, this was a relatively, large study compared to existing literature highlighting the survival benefit that lung transplantation confers to patients with IPF and providing a potential novel biomarker for the pre-lung transplant evaluation of patients with IPF. Lung transplantation substantially prolongs survival of patients with IPF. Incorporation of pre-lung transplant monocyte counts in the pre-transplant evaluation of patients with IPF could optimize the selection of ideal lung transplant candidates with increased probability of survival and identify patients with increased risk of adverse events post lung transplantation. Future prospective studies and larger cohorts of patients will be required before translating our findings into clinical practice.

## Declarations

**Competing interests:** None to declare.

**Funding:** This study was funded by the Ubben Family Fund (JHM).

**Data availability:** Data are available upon request to the corresponding author.

**Ethics approval:** Ethical approval for this study was given by the Institutional Review Boards / Research Integrity & Compliance of University of South Florida (STUDY: 008038).

**Authors approval:** All authors approved this form of the manuscript.

## Data Availability

Data are available upon request to the corresponding author.

## References

1. Navaratnam V, Hubbard RB. The Mortality Burden of Idiopathic Pulmonary Fibrosis in the United Kingdom. American Journal of Respiratory and Critical Care Medicine. 2019;200(2):256–8.

2. Middleton PG, Mall MA, Dřevínek P, Lands LC, McKone EF, Polineni D, et al. Elexacaftor-Tezacaftor-Ivacaftor for Cystic Fibrosis with a Single Phe508del Allele. N Engl J Med. 2019;381(19):1809–19.

3. Chambers DC, Cherikh WS, Harhay MO, Hayes D, Jr., Hsich E, Khush KK, et al. The International Thoracic Organ Transplant Registry of the International Society for Heart and Lung Transplantation: Thirty-sixth adult lung and heart-lung transplantation Report-2019; Focus theme: Donor and recipient size match. J Heart Lung Transplant. 2019;38(10):1042–55.

4. Valapour M, Lehr CJ, Skeans MA, Smith JM, Uccellini K, Goff R, et al. OPTN/SRTR 2018 Annual Data Report: Lung. Am J Transplant. 2020;20 Suppl s1:427–508.

5. George PM, Patterson CM, Reed AK, Thillai M. Lung transplantation for idiopathic pulmonary fibrosis. Lancet Respir Med. 2019;7(3):271–82.

6. Leong SW, Bos S, Lordan JL, Nair A, Fisher AJ, Meachery G. Lung transplantation for interstitial lung disease: evolution over three decades. BMJ Open Respir Res. 2023;10(1).

7. Thabut G, Mal H, Castier Y, Groussard O, Brugiere O, Marrash-Chahla R, et al. Survival benefit of lung transplantation for patients with idiopathic pulmonary fibrosis. J Thorac Cardiovasc Surg. 2003;126(2):469–75.

8. Kapnadak SG, Raghu G. Lung transplantation for interstitial lung disease. Eur Respir Rev. 2021;30(161).

9. Riddell P, Kleinerova J, Eaton D, Healy DG, Javadpour H, McCarthy JF, et al. Meaningful survival benefit for single lung transplantation in idiopathic pulmonary fibrosis patients over 65 years of age. Eur Respir J. 2020;56(1).

10. Swaminathan AC, Hellkamp AS, Neely ML, Bender S, Paoletti L, White ES, et al. Disparities in Lung Transplant among Patients with Idiopathic Pulmonary Fibrosis: An Analysis of the IPF-PRO Registry. Ann Am Thorac Soc. 2022;19(6):981–90.

11. Herazo-Maya JD, Noth I, Duncan SR, Kim S, Ma SF, Tseng GC, et al. Peripheral blood mononuclear cell gene expression profiles predict poor outcome in idiopathic pulmonary fibrosis. Sci Transl Med. 2013;5(205):205ra136.

12. Herazo-Maya JD, Sun J, Molyneaux PL, Li Q, Villalba JA, Tzouvelekis A, et al. Validation of a 52-gene risk profile for outcome prediction in patients with idiopathic pulmonary fibrosis: an international, multicentre, cohort study. Lancet Respir Med. 2017;5(11):857–68.

13. Scott MKD, Quinn K, Li Q, Carroll R, Warsinske H, Vallania F, et al. Increased monocyte count as a cellular biomarker for poor outcomes in fibrotic diseases: a retrospective, multicentre cohort study. Lancet Respir Med. 2019;7(6):497–508.

14. Kreuter M, Lee JS, Tzouvelekis A, Oldham JM, Molyneaux PL, Weycker D, et al. Monocyte Count as a Prognostic Biomarker in Patients with Idiopathic Pulmonary Fibrosis. Am J Respir Crit Care Med. 2021;204(1):74–81.

15. Karampitsakos T, Torrisi S, Antoniou K, Manali E, Korbila I, Papaioannou O, et al. Increased monocyte count and red cell distribution width as prognostic biomarkers in patients with Idiopathic Pulmonary Fibrosis. Respir Res. 2021;22(1):140.

16. Raghu G, Collard HR, Egan JJ, Martinez FJ, Behr J, Brown KK, et al. An official ATS/ERS/JRS/ALAT statement: idiopathic pulmonary fibrosis: evidence-based guidelines for diagnosis and management. Am J Respir Crit Care Med. 2011;183(6):788–824.

17. Raghu G, Remy-Jardin M, Myers JL, Richeldi L, Ryerson CJ, Lederer DJ, et al. Diagnosis of Idiopathic Pulmonary Fibrosis. An Official ATS/ERS/JRS/ALAT Clinical Practice Guideline. Am J Respir Crit Care Med. 2018;198(5):e44–e68.

18. Ley B, Ryerson CJ, Vittinghoff E, Ryu JH, Tomassetti S, Lee JS, et al. A multidimensional index and staging system for idiopathic pulmonary fibrosis. Ann Intern Med. 2012;156(10):684–91.

19. Van Raemdonck DE, Keshavjee S, Levvey B, Cherikh WS, Snell G, Erasmus ME, et al. 5- Year Results from the ISHLT DCD Lung Transplant Registry Confirm Excellent Recipient Survival from Donation after Circulatory Death Donors. The Journal of Heart and Lung Transplantation. 2019;38(4):S103.

20. Mukai S, Hirama T, Onodera K, Watanabe T, Tasaka S, Okada Y. Key predictors of long-term survival after lung transplantation in Japan. Respiratory Investigation. 2025;63(3):265–72.

21. Bos S, Vos R, Van Raemdonck DE, Verleden GM. Survival in adult lung transplantation: where are we in 2020? Curr Opin Organ Transplant. 2020;25(3):268–73.

22. Meyers BF, Lynch JP, Trulock EP, Guthrie T, Cooper JD, Patterson GA. Single versus bilateral lung transplantation for idiopathic pulmonary fibrosis: a ten-year institutional experience. J Thorac Cardiovasc Surg. 2000;120(1):99–107.

23. Vock DM, Durheim MT, Tsuang WM, Finlen Copeland CA, Tsiatis AA, Davidian M, et al. Survival Benefit of Lung Transplantation in the Modern Era of Lung Allocation. Ann Am Thorac Soc. 2017;14(2):172–81.

24. Glanville AR. COUNTERPOINT: Should Every Patient With Idiopathic Pulmonary Fibrosis Be Referred for Transplant Evaluation? No. CHEST. 2020;157(6):1413–4.

25. Leard LE, Holm AM, Valapour M, Glanville AR, Attawar S, Aversa M, et al. Consensus document for the selection of lung transplant candidates: An update from the International Society for Heart and Lung Transplantation. J Heart Lung Transplant. 2021;40(11):1349–79.

26. Mason DP, Brizzio ME, Alster JM, McNeill AM, Murthy SC, Budev MM, et al. Lung Transplantation for Idiopathic Pulmonary Fibrosis. The Annals of Thoracic Surgery. 2007;84(4):1121–8.

27. Karampitsakos T, Juan-Guardela BM, Tzouvelekis A, Herazo-Maya JD. Precision medicine advances in idiopathic pulmonary fibrosis. EBioMedicine. 2023;95:104766.

28. Karampitsakos T, Tourki B, Herazo-Maya JD. The dawn of precision medicine in fibrotic interstitial lung disease. Chest. 2024.

29. Juan Guardela BM, Sun J, Zhang T, Xu B, Balnis J, Huang Y, et al. 50-gene risk profiles in peripheral blood predict COVID-19 outcomes: A retrospective, multicenter cohort study. EBioMedicine. 2021;69:103439.

30. Karampitsakos T, Tourki B, Jia M, Perrot CY, Visinescu B, Zhao A, et al. The transcriptome of CD14+CD163-HLA-DRlow monocytes predicts mortality in Idiopathic Pulmonary Fibrosis. medRxiv. 2024:2024.08.07.24311386.

31. Perrot CY, Karampitsakos T, Herazo-Maya JD. Monocytes and Macrophages: Emerging Mechanisms and Novel Therapeutic Targets in Pulmonary Fibrosis. Am J Physiol Cell Physiol. 2023.

32. Perrot CY, Karampitsakos T, Unterman A, Adams T, Marlin K, Arsenault A, et al. Mast-cell expressed membrane protein-1 is expressed in classical monocytes and alveolar macrophages in idiopathic pulmonary fibrosis and regulates cell chemotaxis, adhesion, and migration in a TGFβ-dependent manner. American Journal of Physiology-Cell Physiology. 2024;326(3):C964–C77.

33. Unterman A, Zhao AY, Neumark N, Schupp JC, Ahangari F, Cosme C, Jr., et al. Single-Cell Profiling Reveals Immune Aberrations in Progressive Idiopathic Pulmonary Fibrosis. Am J Respir Crit Care Med. 2024.

34. Balestro E, Cocconcelli E, Tinè M, Biondini D, Faccioli E, Saetta M, et al. Idiopathic Pulmonary Fibrosis and Lung Transplantation: When it is Feasible. Medicina (Kaunas). 2019;55(10).

35. Schaffer JM, Singh SK, Reitz BA, Zamanian RT, Mallidi HR. Single- vs Double-Lung Transplantation in Patients With Chronic Obstructive Pulmonary Disease and Idiopathic Pulmonary Fibrosis Since the Implementation of Lung Allocation Based on Medical Need. JAMA. 2015;313(9):936–48.

36. Newton CA, Kozlitina J, Lines JR, Kaza V, Torres F, Garcia CK. Telomere length in patients with pulmonary fibrosis associated with chronic lung allograft dysfunction and post-lung transplantation survival. J Heart Lung Transplant. 2017;36(8):845–53.

37. Alder JK, Sutton RM, Iasella CJ, Nouraie M, Koshy R, Hannan SJ, et al. Lung transplantation for idiopathic pulmonary fibrosis enriches for individuals with telomere-mediated disease. The Journal of Heart and Lung Transplantation. 2022;41(5):654–63.

